# Findings of a feasibility study of pre-operative pulmonary rehabilitation to reduce post-operative pulmonary complications in people with chronic obstructive pulmonary disease scheduled for major abdominal surgery

**DOI:** 10.1101/19007914

**Authors:** Lucy L Marlow, Angeline HY Lee, Emma Hedley, Michael P Grocott, Michael C Steiner, J. Duncan Young, Najib M Rahman, Christopher P Snowden, Kyle T S Pattinson

## Abstract

**Background:** Patients with chronic obstructive pulmonary disease (COPD) are at increased risk of complications and death following surgery. Pulmonary complications are particularly prominent. Pulmonary rehabilitation is a course of physical exercise and education that helps people with COPD manage their condition. Although proven to improve health outcomes in patients with stable COPD, it has never been formally tested as a pre-surgical intervention in patients scheduled for non-cardiothoracic surgery. If a beneficial effect were to be demonstrated, pulmonary rehabilitation for pre-surgical patients with COPD might be rapidly implemented across the National Health Service, as pulmonary rehabilitation courses are already well established across much of the United Kingdom (UK).

**Methods:** We performed a feasibility study to test study procedures and barriers to identification and recruitment to a randomised controlled trial testing whether pulmonary rehabilitation, delivered before major abdominal surgery in a population of people with COPD, would reduce the incidence of post-operative pulmonary complications. This study was run in two UK centres (Oxford and Newcastle upon Tyne).

**Results:** We determined that a full randomised controlled trial would not be feasible, due to failure to identify and recruit participants. We identified an unmet need to identify more effectively patients with COPD earlier in the surgical pathway. Service evaluations suggested that barriers to identification and recruitment would likely be the same across other UK hospitals.

**Conclusions:** Although pulmonary rehabilitation is a potentially beneficial intervention to prevent post-operative pulmonary complications, a randomised controlled trial is unlikely to recruit sufficient participants to answer our study question conclusively at the present time, when spirometry is not automatically conducted in all patients planned for surgery. As pulmonary rehabilitation is a recommended treatment for all people with COPD, alternative study methods combined with earlier identification of candidate patients in the surgical pathway should be considered.

**Trial registration:** ISRCTN 29696295, http://www.isrctn.com/ISRCTN29696295, registered 31st August 2017

## Introduction

In the United Kingdom, chronic obstructive pulmonary disease (COPD) affects approximately 3.7 million people (1), is responsible for approximately 30,000 deaths per year, and is the fifth most common cause of death (2). COPD is an independent risk factor for postoperative complications (odds ratio OR 1.35 (CI 1.30-1.40)) and death (OR 1.29 (CI 1.19-1.39))(3-6). Complications include pulmonary and cardiac events, sepsis, renal insufficiency and an increased reoperation rate (3). Surgical patients with COPD thus represent a high-risk group in whom there is an unmet need to improve post-operative outcomes.

### Pulmonary rehabilitation

Pulmonary rehabilitation is “*A physical exercise and education programme, tailored for each person. It includes information on looking after the body and lungs, advice on managing symptoms, including feeling short of breath, nutrition and psychological support. People who smoke are given advice on how to stop*.*”* (7)

Pulmonary rehabilitation is usually delivered in an outpatient setting, consisting of one hour of exercise and one hour of education, twice weekly for six weeks. It has profound benefits on breathlessness, exercise capacity and quality of life (number needed to treat (NNT)=2) (8), no side effects are reported (9). Pulmonary rehabilitation is associated with decreased hospital admissions (NNT=3-4), and mortality (NNT =∼6) following COPD exacerbations (10-13). Crucially, pulmonary rehabilitation is inexpensive (9). Its effect is so powerful that it has a negative cost per quality adjusted life year (QALY), meaning it saves money for the NHS (14). The main challenges facing pulmonary rehabilitation are the barriers to its uptake as attendance and completion of the programme is often poor (15, 16).

### Improving post-operative outcomes

Despite adoption in NICE guidelines (17) for stable COPD, pulmonary rehabilitation is not regularly offered to pre-surgical patients with COPD(18). We believe that pulmonary rehabilitation merits investigation as a potential means to improve postoperative outcome in people with COPD undergoing surgery for the following reasons:

- A handful of small surgical studies suggest beneficial effects of pulmonary rehabilitation on the incidence of postoperative pulmonary complications (19-23). Differing endpoints, small sample sizes and restriction to specific surgical groups limits conclusive interpretation.
- In the National Emphysema Treatment Trial (NETT) (24, 25), lung volume reduction surgery was compared with medical management of COPD. All patients underwent pre-operative pulmonary rehabilitation. In the thoracic surgical population of NETT similar outcomes (in terms of functional exercise capacity and health related quality of life, assessed prior to surgery) were observed to what would be expected in the treatment of non-surgical patients with COPD. In fact some participants in NETT decided against lung volume reduction surgery because they felt so much better after pulmonary rehabilitation.
- Shortened durations of pulmonary rehabilitation are efficacious (26, 27). This is important, because an adapted course may be necessary to fit within surgical time frames.
- Pulmonary rehabilitation is widely available and standardised across the NHS in over 200 UK centres. This has important implications for scalability.

Pre-operative pulmonary rehabilitation needs sufficient time between the decision to operate and the operation, requires cross specialty working, and involves patients with two conditions (COPD and a surgical condition). A randomised controlled trial is therefore justified, as the current evidence base is either not specific to a surgical population or is case series based and therefore subject to selection bias. Furthermore, it is unclear whether a randomised controlled trial of pulmonary rehabilitation before surgery would be practical. This study investigated the feasibility of running such a large randomised controlled trial. The feasibility study design matched the expected full study design except in scale.

## Methods

This feasibility study was conducted across two research sites (Oxford and Newcastle upon Tyne), chosen as two areas with different demographics and incidence rates of COPD. Ethical approval was granted by the South Yorkshire Research Ethics Committee (approval number 17/YH/0220). Written informed consent was obtained from all participants prior to the start of the study. The primary aim of the study was to determine feasibility for a randomised controlled trial and focused on recruitment rate, barriers to recruitment and uptake of pulmonary rehabilitation.

### Study procedures

The main inclusion criteria were

1. Patients scheduled for elective major (body cavity) surgery excluding cardiothoracic and orthopaedic surgery. In practical terms this meant that recruitment focused upon patients undergoing surgery for abdominal cancer.
2. A diagnosis of COPD. COPD was defined as post bronchodilator FEV1/FVC<0.70, FEV1≤ 80% normal. Patients with a physician diagnosis of COPD were included even if spirometry was not immediately available.

The main exclusion criterion was patients who were unable to participate in pulmonary rehabilitation according to the British Thoracic Society guidelines (28). Full inclusion and exclusion criteria can be found in the appendix.

### Participant identification and recruitment

To determine the best point in the surgical pathway to recruit participants, research nurses tested feasibility of screening for study participants from the following sources.

- From the surgical multidisciplinary team (MDT) meetings
- In oncology clinics
- From the electronic patient record for patients scheduled for surgery
- From hospital anaesthetic preoperative assessment clinics
- From cardiopulmonary exercise testing clinics

The study aimed to collect 48 full data sets (24 in each centre, 12 pulmonary rehabilitation, 12 control arm). To achieve this 48 dataset target, based on known drop-out rates from pulmonary rehabilitation (18) and potential further data loss due to surgical scheduling, we anticipated that we would need to recruit 72 patients.

### Pulmonary rehabilitation

A pragmatic, exploratory approach was used to explore what is practically deliverable and tolerated by patients, working closely with local pulmonary rehabilitation teams in Oxford and Newcastle upon Tyne. The aim was for patients to be enrolled in 3 pulmonary rehabilitation sessions per week, for 3 or 4 weeks, depending on timing of surgery. Pulmonary rehabilitation of this shortened duration has been shown to be effective(27). Patients were to attend standard NHS pulmonary rehabilitation groups run for patients with COPD.

### Control arm

Patients randomised to the control arm would receive standard care including advice on smoking cessation, exercise and appropriate referral and education for those with newly diagnosed COPD.

**Research assessments** are described in full in the Appendix. Randomisation was 1:1 to either pulmonary rehabilitation or control with self-report questionnaires and clinical outcome scores collected during the hospital stay and with a 6-week and 6-month follow up.

**Feasibility measures** collected details of barriers to participant identification, recruitment and retention, the demographics of recruited participants and the feasibility of delivering pulmonary rehabilitation in the time available. We assessed study logistics, performance of research measures, the effectiveness of randomisation. The full list of feasibility measures is detailed in the appendix.

### Outcome measures

We anticipated that the primary research outcome measures for a future randomised controlled trial would be morbidity, mortality, length of hospital stay and hospital readmissions so we collected data on this to help with future study design.

## Results

Recruitment commenced in Oxford in January 2018 and in Newcastle upon Tyne in May 2018. A total of 266 patients were screened of which 65 met the inclusion criteria. As of January 2019, one participant had been recruited in Oxford and two in Newcastle upon Tyne. At this point it was determined that running a randomised controlled trial of pulmonary rehabilitation would not be feasible and the study was terminated. Further details are presented in Figure 1. We have not presented the research data here due to interpretability and confidentiality issues arising from only acquiring two datasets.

**Figure 1:**
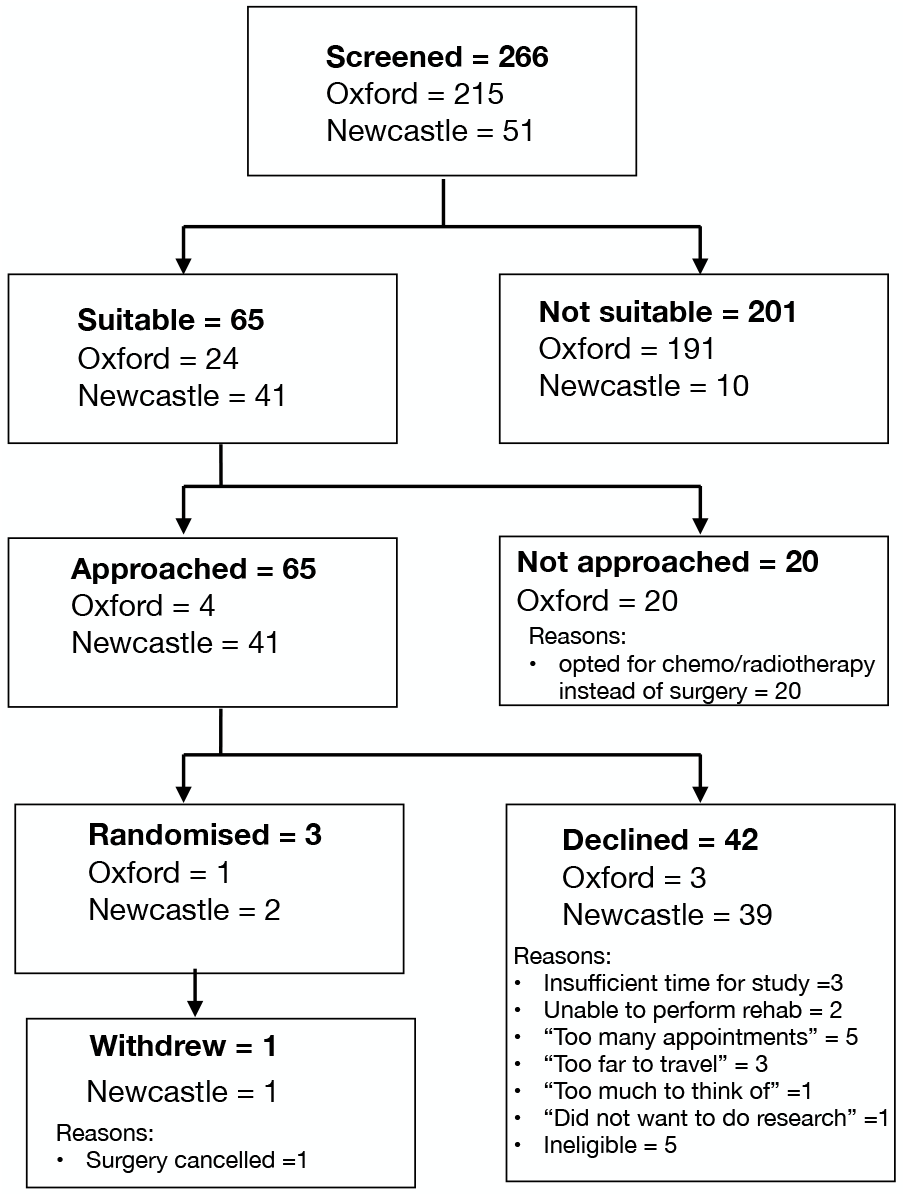
Flow diagram of participant identification and recruitment

We found that the main barrier to study recruitment in both centres was associated with the way the surgical pathway is organised, especially with regards to two specific aspects; surgical timelines and identification COPD.

### Barriers to identification of study participants

In Oxford, challenges were faced in identifying patients with COPD soon enough before surgery.

It was challenging to identify patients with COPD at surgical clinics and multidisciplinary meetings as patients had just received a diagnosis of cancer but a definitive treatment plan had yet to be instituted. At this point the focus is on the surgical condition rather than medical conditions such as COPD. Medical records focused mostly upon surgical condition and respiratory records were often in separate (unavailable) notes and smoking histories were rarely present. This made screening laborious and time inefficient.

The definitive decision on whether to operate would only be made following neoadjuvant treatment. Oncology clinics were assessed as an identification point, but we found that potential participants attended too many different clinics to find a suitable point for screening.

Screening the electronic patient record for patients scheduled for surgery did not successfully identify additional people with COPD. Therefore, COPD was often not formally diagnosed until the following the anaesthetic preoperative assessment clinic which usually occurred 2-3 weeks before surgery, with CPET testing taking place a similar time before surgery.

The difficulty in identifying potential participants with COPD was somewhat unexpected. As audit data from the pre-operative cardiopulmonary exercise testing clinics in both Oxford and Newcastle-upon-Tyne suggested that COPD was present in 10 to 15% of the 2,000 to 3,000 patients each year passing through those clinics. This meant that our pool of potential participants was around 300 in each centre each year.

Vascular surgery clinics were also assessed; these non-cancer patients have a more clearly defined pre-surgical pathway. However, we found that due to changes in surgical practices, most patients with respiratory disease were treated endovascularly and thus recruiting from this clinic was also deemed low yield.

At anaesthetic pre-assessment clinics, the main challenge was that potential participants with undiagnosed COPD may not have been formally diagnosed after the pre-assessment clinic (when patients were sent for lung function tests); this made confirmation of eligibility difficult, and further lessened time for study inclusion.

### Barriers to recruitment of study participants

In Newcastle upon Tyne, surgical patients attend the anaesthetic pre-assessment clinic about one month before surgery, this is in contrast to Oxford where the time between anaesthetic assessment and surgery is often much shorter. In Newcastle-upon-Tyne we were more successful at identifying patients with COPD, but despite this only two patients were recruited into the study (one of whom subsequently withdrew).

### National survey of preassessment clinics

We discussed increasing the number of sites for the study with three other potential UK sites (two teaching hospitals and one large district general hospital) who performed evaluations of their services, taking into account the preliminary findings of this work. This would help us evaluate whether the identification and recruitment issues were generalisable to other centres. However, we found that in all three centres main point of identifying COPD was found to be at anaesthetic pre-assessment clinics, which occur two to three weeks prior to operation date (similar to Oxford).

CPS, in his role as Royal College of Anaesthetists National Clinical Lead for Perioperative Medicine, surveyed perioperative medicine and preoperative assessment clinics in 110 hospitals in England over the course of 2017 (29). This piece of work found that the usual time interval between anaesthetic preassessment and surgery was often only 2-3 weeks, but with wide variability (unpublished observations). This is equivalent to current practice in Oxford.

## Discussion

Pulmonary rehabilitation is a potentially valuable treatment for improving the health status of people with COPD prior to surgery. We established that a full randomised controlled trial is not feasible. As a result of this study we have identified an unmet need in the early identification of COPD in patients presenting for surgery.

Although the study was only run in two UK centres, further scoping work in three additional centres and a related England-wide survey of anaesthetic services means that we are reasonably confident that similar challenges in identification and recruitment would be found if a randomised controlled trial were to run across the UK. However, on-going developments in the reorganisation of perioperative medicine services, such as joint surgical and anaesthetic clinics and longer time intervals between anaesthetic assessment and surgery in the medium term (next 5 years) such trial may be possible.

Pulmonary rehabilitation is an integral part of the NICE guidelines for the treatment of COPD, and therefore every person with COPD should be offered this treatment (alongside the other components of therapy recommended by NICE). This raises the question about whether a randomised controlled trial is actually the most appropriate methodology for future work. Barriers to pulmonary rehabilitation are well recognised, even when implemented a clinical treatment (15, 16). These barriers can be even more pronounced when tested as an optional research intervention (30). We provided free transport and offered flexible scheduling for potential participants. These were recommended by our patient liaison group during the study design phase to help overcome barriers to taking part in pulmonary rehabilitation, but clearly were insufficient to enable us to recruit at a sufficient rate.

We therefore speculate that if it we could identify COPD at the beginning of the patient’s surgical journey, patients would be much better placed to have appropriate management and optimisation of their COPD. Spirometry is cheap, widely available and reliable and thus perfect for a simple primary care test which should be offered much more widely and would allow for early optimisation of drug therapy. Pulmonary rehabilitation could occur in a more timely fashion as this intervention could be integrated and planned alongside chemo- and radio-therapy, rather than in the weeks immediately preceding surgery. Patients with COPD would benefit even if they do not eventually proceed to surgery, with potential cost savings for the NHS(34, 35). Potential solutions to this are illustrated in Figure 2.

**Figure 2.**
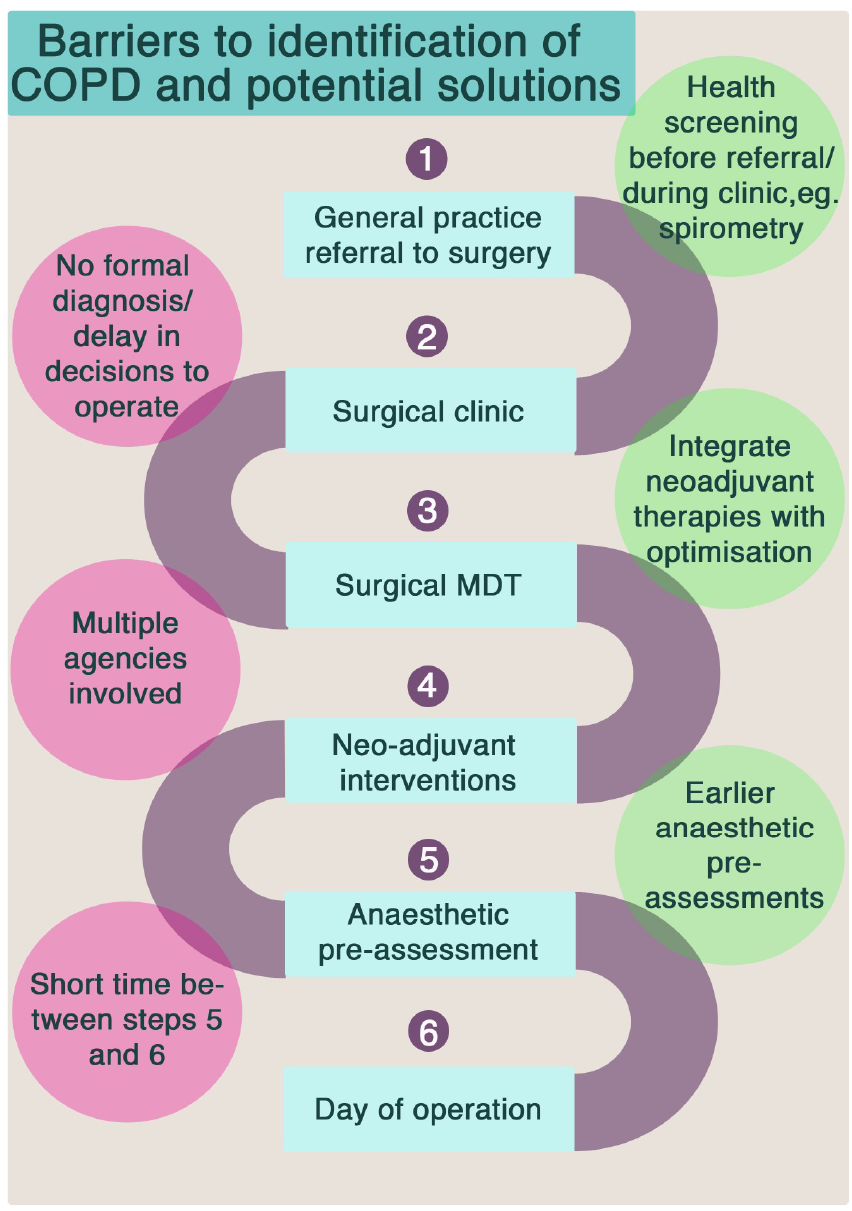
A summary of the challenges to identification (and thus treatment) of COPD for surgical patients and potential solutions

This study has demonstrated the considerable challenge in performing additional interventions in the immediate period before surgery. However, the duration of the patient’s journey from referral to surgery can take several months and remains an ideal period to optimise COPD if appropriate patients are identified earlier in the process. Some UK hospitals have recently implemented initiatives to ‘re-design’ this surgical pathway, which may help overcome this barrier (36). The importance of identifying and engaging with patients early after the “moment of contemplation” of surgery is clearly a critical success factor for interventions such as pulmonary rehabilitation; which are known (from other contexts) to require a defined period of time to implement and provide benefit. However, we should take caution from evidence from studies in lung cancer which show that the time of diagnosis is a difficult time to consider pulmonary rehabilitation (31). Although there may be an opportunity to provide pulmonary rehabilitation whilst neo-adjuvant therapy is being provided patients often do not want to engage with pulmonary rehabilitation at a time when they are dealing with a new, life changing diagnosis and having burdensome, potentially toxic cancer treatment (31). Thus it might turn out that pulmonary rehabilitation can only really feasibly delivered once cancer treatment has finished. There is emerging evidence that exercise therapies enhance cancer survival (32, 33) and that a recommendation from the oncologist may be influential in the view patients might take.

Pulmonary rehabilitation represents an important part of the NICE guidelines for the treatment of COPD and is readily available in the NHS. We know that patients will benefit from pulmonary rehabilitation, even if it is consequently shown not to have a specific effect upon postoperative pulmonary complications. Patients who consequently do not require surgery will still benefit. We have shown that a randomised controlled trial is not feasible, so we need to approach this in a different way using alternate methodologies.

## Data Availability

not applicable

## Declarations

### Ethics approval and consent to participate

Ethical approval was granted by the South Yorkshire Research Ethics Committee (approval number 17/YH/0220). Written informed consent was obtained from all participants prior to the start of the study.

### Consent for publication

Not applicable

### Availability of data and material

Not applicable

### Trial registration

ISRCTN 29696295, http://www.isrctn.com/ISRCTN29696295, registered 31st August 2017

### Authors’ contributions

LM: Contributed to data collection and pathway appraisal, and drafted the manuscript. Contributed and approved the final manuscript.

AL: Contributed to the design of the research question and contributed and approved the final manuscript

EH: Contributed to the design of the research question and study assessments, and contributed and approved the final manuscript

MG, MS, JDY, NR, CS: Conceived and designed the research question, and contributed and approved the final manuscript

KP: Conceived and designed the research question. Contributed to data collection and pathway appraisal, and drafted the manuscript. Contributed and approved the final manuscript.

## Appendix

### Patient involvement in study design

As we anticipated that recruitment to this study may be challenging, we discussed the study design with patient groups consisting of people with COPD who had either undergone surgery, or those who had experienced pulmonary rehabilitation. The key messages from these patient representatives were to ensure that transport to and from pulmonary rehabilitation would be provided, and that a flexible approach to scheduling would be necessary so pulmonary rehabilitation could fit with other appointments.

### Inclusion Criteria

- Adult patients aged 18 years or older with COPD
- Has capacity to take part in this study
- Scheduled for elective major (body cavity) surgery OR laparascopic surgery
- anticipated to last longer than 2 hours
- People with more than 20 pack years smoking history were approached to take part in the study if spirometry subsequently confirmed COPD.

### Exclusion Criteria

- Inability to give informed consent
- Insufficient command of English to understand the study documentation
- Unable to participate in pulmonary rehabilitation treatment according to British
- Thoracic Society guidelines.
- Patients scheduled cardiac, thoracic and orthopaedic surgery and orthopaedic surgery

---

**Figure 1.**
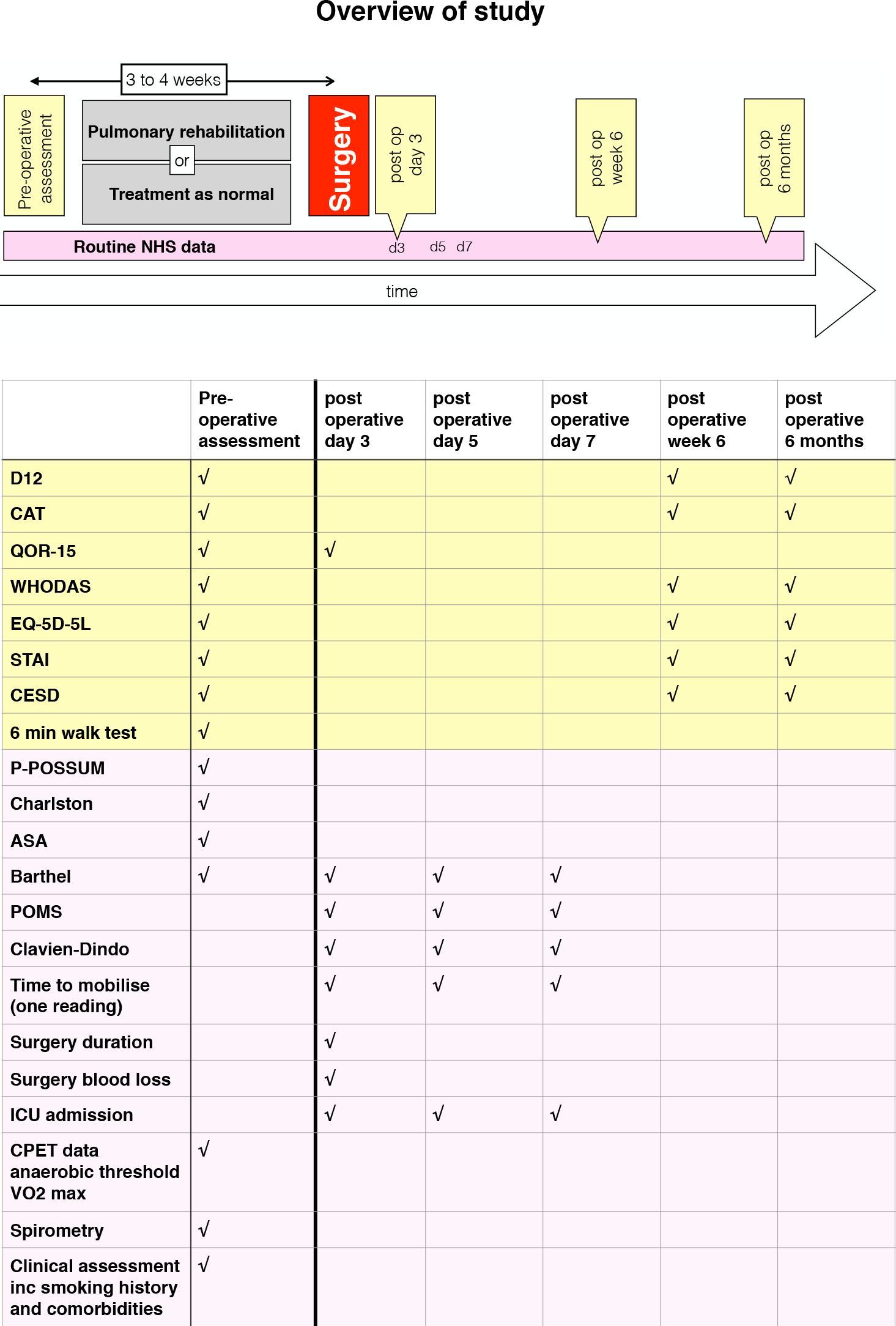
Overview of study methods. Data in yellow boxes is research data collected from the patient, whereas the data in pink relates to that collected from the patients’ clinical record. Abbreviations: NHS; National Health Service, D12; Dyspnoea-12 questionnaire, CAT; COPD assessment test, QOR-15; quality of recovery score, WHODAS; World Health Organisation (WHO) disability score, STAI; Spielberger state and trait anxiety inventory, CESD; Center for epidemiolgic studies depression scale, P-POSSUM; Portsmouth Physiological and Operative Severity Score for the enumeration of Mortality, Charlston; Charlston Morbidity Index, ASA; American Society of Anesthesiologists Physical Status Classification System, Barthel; Barthel scale, Clavien-Dindo; The Clavien-Dindo Classification of surgical complications, ICU; intensive care unit, CPET; cardiopulmonary exercise test.

## Feasibility measures collected throughout the study

### Key feasibility measures

- Can we recruit at a sufficient rate to run an RCT?
- What is severity (GOLD/MRC) of the recruited patients and how does this compare with the screened patients
- Whether it is feasible to deliver the pulmonary rehabilitation intervention in the time available. This will include assess the impact of changing surgical dates, e.g. earlier (so insufficient rehabilitation delivered), later (so effect of rehabilitation wearing off).
- Number of complete data sets collected
- %missing data
- Barriers to uptake of pulmonary rehabilitation

### We will detail recruitment, retention and dropouts (including a screening log) at all the potential drop out points

- Number of patients identified in clinic with spirometry defined COPD, and their MRC and GOLD scores
- Number of patients invited to participate in the study, and their MRC and GOLD scores
- Number who accept invitation
- Number who decline invitation but agree to participate in qualitative study
- Number who decline invitation/don’t reply
- Number of patients who attend research assessment
- Number of patients who sign consent form
- Number of patients who complete pulmonary rehab or control treatment (i.e. compliance with study intervention)
- Number of patients who have surgery in allocated timeframe (<3/12followingresearchvisit2)
- Number of patients who continue the study during postoperative period
- Number of patients in whom we can collect 6-month follow up data.

### Logistics. We will collect measures relating to

- Scheduling of research appointments within suitable timef rames
- Scheduling of pulmonary rehabilitation sessions within the surgical waiting time
- Effectiveness of transport to/from pulmonary rehabilitation. Although we plan to contribute transport costs for the study there needs to be consideration for when pulmonary rehabilitation is offered as a treatment.
- Factors relating to scheduling, including effect of changes in operation date.
- Feasibility of tracking patients postoperatively-either in person and/or via electronic and paper based patient records

### Performance of measures including ceiling and floor effects Outcome measures being collected to get an estimate of

- central tendency
- spread
- data loss
- loss to follow up
- Event rate of postoperative complications, to help with sample size calculation for main study

### Effectiveness of randomisation

- Check for post randomisation drop outs because allocated to unfavoured treatment group
- Do the “treatment as normal” patients seek exercise sessions elsewhere?
- Is the drop-out rate from study similar in both groups?
- Does the randomisation system work?

### Health economics

We know there is health economic benefit for pulmonary rehabilitation in the treatment of COPD -does this translate to a surgical population?

- EQ-5D-5L measured at baseline, at day 5, 6 weeks and 6 months post operatively
- Resource use will be measured, including primary care, pulmonary rehabilitation, hospital services(e.g. before during and after surgery)

### Plans to mitigate against bias / Outcome integrity

- Assessors will be blinded to treatment group.
- We will trial ways to ensure that the outcomes chosen are as fair as possible and are collected in a way that avoids bias. This will include objective criteria scoring by blinded individuals.

## Research assessments

Recruited participants were randomised 1:1 to either pulmonary rehabilitation or treatment as normal, minimised for study site and Global Initiative for Chronic Obstructive Lung Disease (GOLD) stage, using an online randomisation service (Sealed Envelope Limited, London, UK). Participants were assessed prior to pulmonary rehabilitation or control treatment (preoperative assessment) and following surgery during hospital inpatient stay on postoperative days 3, 5, and 8 and again at a 6-week and 6-month follow-up. The data collected at each time point can be found in the appendix. Other than a 6-minute walk test, the research data consisted of self-report questionnaires on mood, symptoms, and quality of life. Data obtained from the NHS clinical record included various perioperative risk scores, co-morbidity scores, and measures relating to the operation and outcomes.

### Preoperative (prior to pulmonary rehabilitation or control treatment)

- Medical, surgical, anaesthetic assessment including comorbidities including full detailed smoking histories.
- Physiology
  - Spirometry
  - 6 minute walk test
  - Cardiopulmonary exercise testing (CPET)
  - Physical activity monitoring (accelerometry-based wristwatch) – monitored for one week.
  - Preoperative risk assessment scoring using POSSUM-R, Charlston Co-morbidity Index, ASA grade.

- Psychology and health-related quality of life
  - Dyspnoea questionnaires (Dyspnea-12 questionnaire)
  - Anxiety (State and Trait Anxiety Inventory), Depression (Center for Epidemiological Studies Depression Scale), Fatigue (Fatigue Severity Scale), COPD Assessment Test (CAT)
  - Health status assessment with EQ-5D-5L and WHO disability assessment schedule.

### Post-operative measures

- These measures will be collected on postoperative days 3, 5, and 8 during hospital inpatient stay.
  - Surgical factors (duration of operation, blood loss), measured once only
  - Time to mobilisation
  - Assessment of activities of daily living (Barthel).
  - Intensive care admission, discharge, mortality
  - Patient-related outcome measures, including time to return to normal activities.

- Morbidity tracking using the postoperative morbidity survey instrument and Clavien-Dindo surgical complication score.
- Postoperative measures collected on day 5 post-surgery only
  - Health status questionnaires: Dyspnea, Anxiety, Depression, CAT EQ-5D-5L and WHO disability assessment schedule
  - Smoking history

- Data collected at discharge from hospital
  - Date of discharge (i.e. length of hospital stay)
  - Destination of discharge

- Postoperative measures collected during 6-week follow-up visit.
  - Health status questionnaires: Dyspnea, Anxiety, Depression, CAT EQ-5D-5L and WHO disability assessment schedule
  - Readmissions to hospital, morbidity tracking as above (from medical record).
  - Smoking history

- Postoperative measures collected 6 months following surgery
Following confirmation that patient remains alive (NHS Spine and communication with general practitioner) we will invite the patient to attend a follow up assessment and collect the following measures.
  - Health status questionnaires: Dyspnea, Anxiety, Depression, CAT EQ-5D-5L and WHO disability assessment schedule
  - Readmissions to hospital, morbidity tracking as above (from medical record).
  - Smoking history

### Pulmonary rehabilitation

This section gives further information on the composition of standard course of pulmonary rehabilitation that is used for the treatment of COPD in the non-surgical setting. The courses are preceeded by an assessment session to check suitability for pulmonary rehabilitation and collect baseline clinical measures (based around functional exercise capacity, quality of life and symptom measures, and spirometry). The final session repeats these assessments.

The courses usually consist of two hour sessions performed twice weekly for six weeks. These consist one hour exercise and one hour education. The exercise sessions include both aerobic and strengthening exercises and are tailored to the individual’s ability and there may be variation between courses.

Aerobic exercises may include:

- Step-ups
- Walking on the spot
- Treadmill walking
- Exercise bicycle

Strengthening exercises (3 sets of 10) include:

- Sit-to-stand
- Biceps curls
- Upright row
- Leg extensions

Education sessions may include

- Introduction to rehabilitation
- Management of breathlessness
- Airway clearance
- Understanding your lung condition
- Home exercise program -adding in your own exercises
- Medicine management
- Staying healthy
- Stress and relaxation
- Pacing and energy conservation
- Healthy diet
- Smoking cessation
- Continuing support
- Advanced care plans
- Sexual function

